# Dysregulation of microRNAs in fibromyalgia: potential as biomarkers and disorder modulators

**DOI:** 10.64898/2026.09.08.26362323

**Authors:** Alessandra Tamashiro-Orrego, David A. Andersson, Rocio T Martinez-Nunez

## Abstract

Fibromyalgia syndrome (FMS) is characterized by chronic widespread pain, fatigue and cognitive difficulties, but its underlying mechanisms remain incompletely understood. The uncertainty of the origins of FMS has hampered the development of treatments and diagnostic methods, and the available therapeutic options offer limited relief to patients, an estimated 2-4% of the population. The severe burden of FMS thus represents an urgent unmet clinical need.

MicroRNAs (miRNAs), non-coding short RNAs that have vital inhibitory roles in gene expression, have been explored in FMS pathophysiology. MicroRNA signatures associated with FMS may facilitate diagnosis and reveal new insights into the cellular and molecular mechanisms of FMS.

We compiled and analyzed publicly available data of dysregulated miRNAs in FMS. MiRNAs were categorized according to the direction of dysregulation (levels up or down in FMS vs health) and compared across studies. Only miRNAs identified as dysregulated in the same direction in at least two studies were included. RNA targets of at least two microRNAs were inferred using TargetScan and subjected to pathway enrichment analysis using Reactome, ShinyGo v.8, miRNA Tissue Atlas 2025 and EnrichR.

This analysis revealed enrichment in pathways central to cell signaling and immune function, such as regulation of PTEN, circadian rhythm and RNA metabolic processes. Additionally, the dysregulated miRNAs in FMS were over-represented in immune-related tissues such as lymph node and spleen.

Collectively, these findings support a role for immune dysregulation in FMS. By using an integrative approach, we identified candidate pathways and molecular targets that warrant further experimental validation.

## Introduction

Fibromyalgia syndrome (FMS) is a chronic primary pain condition associated with a diverse set of symptoms, including widespread pain, persisting fatigue, unrefreshing sleep, and cognitive impairment (Hauser et al., 2015; Nicholas et al., 2019). FMS is a highly prevalent diagnosis globally, affecting 2-4% of the world’s population, predominantly females (Wolfe et al., 2018). Despite its prevalence and considerable impact on the quality of life of patients, the underlying causes of FMS remain incompletely understood.

Numerous neurological and immunological aberrations have been reported in patients with FMS. These include dysregulated neurotransmitter levels (Becker and Schweinhardt, 2012), hyperexcitable C-fiber nociceptors (Serra et al., 2014), small-fibre pathology (Oaklander et al., 2013; Üçeyler et al., 2013), neurovascular abnormalities (Albrecht et al., 2013), elevated pro-inflammatory cytokines/chemokines (Rodriguez-Pinto et al., 2014), structural mitochondrial aberrations (Israel et al., 2024), and an altered microbiome (Cai et al., 2025). Despite the wealth of data demonstrating that FMS diagnosis is associated with peripheral pathophysiology, the condition is widely thought to be caused by aberrant central cortical sensory processing (Nijs et al., 2021). Reverse translational investigations have provided direct support for a causal role of immunoglobulin G (IgG) antibodies in FMS, since transfer of purified patient IgG to mice recapitulated sensory and motor symptoms and signs of small-fibre pathology in mice (Goebel et al., 2022; Goebel et al., 2021). Electrophysiological investigations of mice treated with patient IgG further identified sensory afferent abnormalities that mirror those observed in patients (Israel et al., 2025; Israel et al., 2026). It thus seems likely that FMS symptoms are driven by autoreactive IgG acting systemically.

Diagnosis of FMS is exclusively based on clinical assessment of symptoms, and since it is not a diagnosis of exclusion, FMS is often comorbid with other disorders (Fitzcharles et al., 2018). There is currently no defined clinical biomarker, and the available pharmacotherapies include anti-epileptic and anxiolytic medications such as gabapentinoids, duloxetine, and milnacipran that are thought to mitigate the impact of symptoms by modulating neurotransmitter levels (Giorgi et al., 2024). Patients may also pursue non-pharmaceutical approaches such as gentle, supervised exercise and cognitive behavioral therapy to manage symptoms (Bernardy et al., 2010; Hauser et al., 2010). However, all these approaches, focused on modulating central nervous system processes, have limited efficacy leading to only modest improvements of pain and associated symptoms in most patients (Clarke et al., 2025). The modest efficacy and the high rates of side effects of available drugs, led the UK National Institute for Health and Care Excellence (NICE) to discourage initiation of analgesics (Harvey-Sullivan et al., 2022) for fibromyalgia and other chronic primary pain disorders. Pain management in FMS is therefore an urgent unmet clinical and societal need, and investigations focused on elucidating the mechanisms responsible for FMS are paramount to accelerate the development and introduction of more effective therapies.

In recent years, microRNA (miRNA) signatures have emerged as promising biomarkers for complex diseases in the fields of infection, immunity, cancer and neuroscience (Condrat et al., 2020). MiRNAs are non-coding small RNAs, around 22 nucleotides long, that inhibit gene expression post-transcriptionally. A single microRNA can bind to complementary sequences in the 3’ un-translated regions (3’UTRs) of multiple target messenger RNAs (mRNAs) (Oliveto et al., 2017) and, conversely, one mRNA may be targeted by several microRNAs. MiRNA binding can inhibit mRNA translation into protein, and/or trigger mRNA degradation. Thus, miRNAs are important regulators of cellular homeostasis, and miRNA dysregulation has been linked to a number of pathological conditions (Juzwik et al., 2019).

MiRNAs are stable in biofluids, particularly when contained in extracellular vesicles. Extracellular vesicles are lipid bilayered nano-vesicles released by all cells as a form of communication (Xu et al., 2022). Several studies have investigated and reported altered miRNA expression levels in patients with FMS in a wide variety of tissues, mainly in blood or plasma (Akaslan et al., 2021; Bjersing et al., 2015; Bjersing et al., 2013; Braun et al., 2020; Cerdá-Olmedo et al., 2015; Clos-Garcia et al., 2019; Erbacher et al., 2024; Erbacher et al., 2022; Hernandez et al., 2024; Hussein et al., 2022; Leinders et al., 2016; Masotti et al., 2017; Nepotchatykh et al., 2023; Rasulova et al., 2024). Crucially, beyond their diagnostic use, dysregulated microRNAs may also provide important novel insights into pathophysiological mechanisms (Palanichamy and Rao, 2014).

While multiple studies have identified several dysregulated miRNAs in FMS, these findings have been heterogenous, and the specific miRNAs identified as differentially expressed in FMS have varied between cohorts and were assessed using different methods. Moreover, not all studies emphasized the target, or potential target, mRNAs of the dysregulated miRNAs or the associated dysregulated pathways in patients. This biological information could provide crucial insight into the underlying mechanisms of FMS, the role of miRNAs in FMS and/or their potential use to understand underlying disease mechanisms.

Here, we analyze all publicly available data from studies of miRNA expression in human samples from FMS patients. We identified miRNAs reported as dysregulated in at least two distinct studies and perform bioinformatics analyses to infer potentially affected molecular pathways that can inform us about FMS mechanisms. By curating existing data in this manner, we aimed to highlight regulatory networks that might warrant prioritization in future experimental investigations.

## Methods

### Literature Search

PubMed (https://pubmed.ncbi.nlm.nih.gov/) was used with the search terms "microRNA" or "miRNA" and "fibromyalgia" or “FMS” and included studies published up to October 2025. This search yielded 40 results. We included primary research studies based on human samples that also provided a list of significantly dysregulated miRNAs (if any). 15 of the 40 studies met the criteria. 14 reported changed levels of miRNAs in FMS (Figure 1), and from those, the lists of dysregulated miRNAs were extracted.

**Figure 1.**
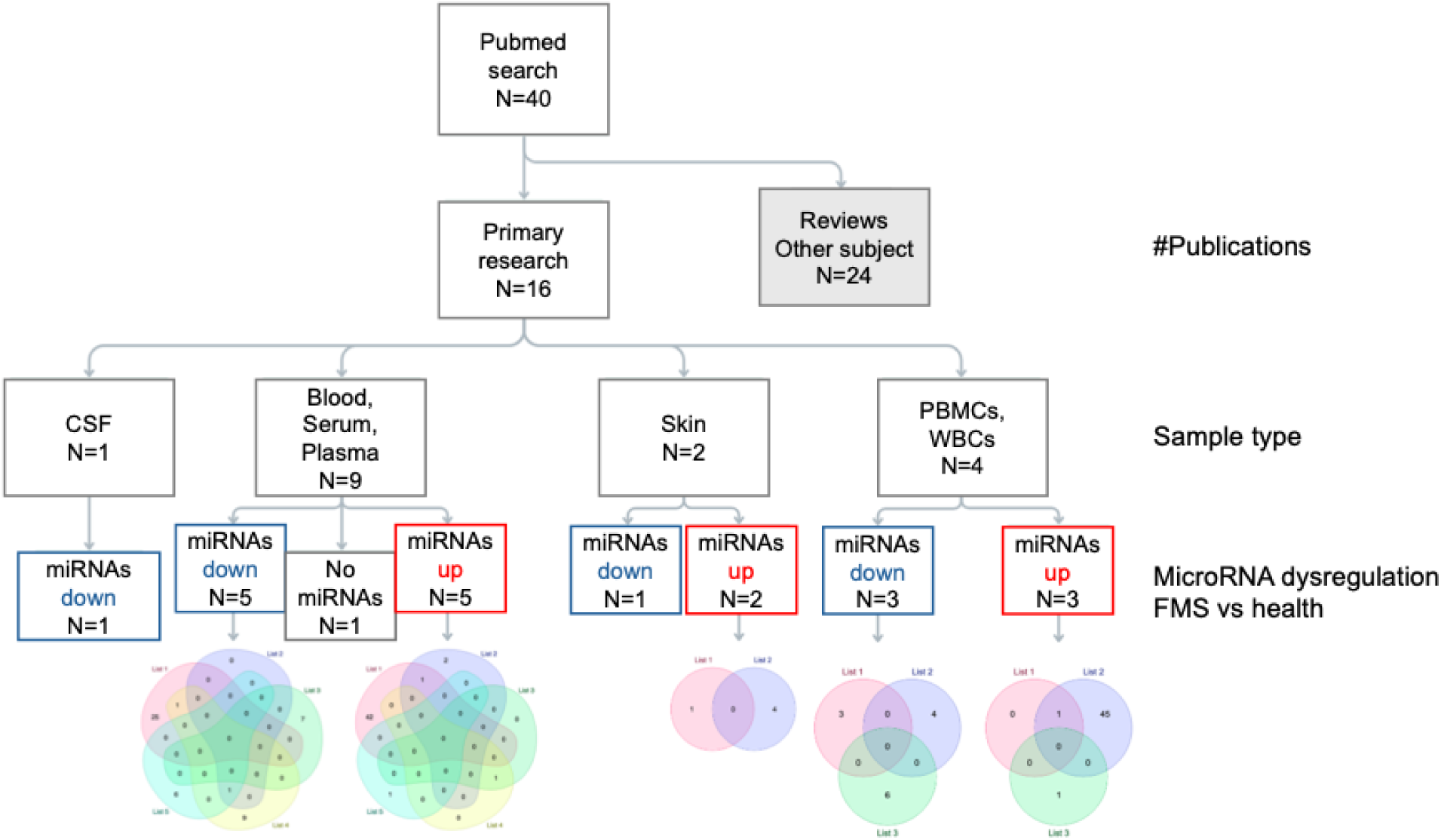
Flow diagram of the approach to find commonly dysregulated microRNAs in FMS patients using public datasets. Literature search was conducted to find primary research studies that had discovered dysregulated levels of miRNAs in FMS patients. CSF: cerebrospinal fluid; PBMCs: peripheral blood mononuclear cells; WBS: whole blood cells.

When necessary, miRNA nomenclature was standardized according to miRbase registry (Kozomara et al., 2019). This included consolidating the old nomenclature with the up-to-date -3p and -5p denominations. Following this, miRNA lists were compared to identify common patterns of microRNA dysregulation.

### MicroRNAs Target Prediction

MicroRNAs reported as dysregulated in two or more studies were selected for downstream analysis and categorized based on the direction of the changes in their levels (downregulated or upregulated relative to controls). Target prediction was performed using TargetScan v8.0 (McGeary et al., 2019) for each identified human miRNA, with targets ranked according to weighted context++ score. Targets with a weighted context++ score of -0.2 or less were extracted and used for subsequent pathway analysis.

### Pathway Prediction Analysis

Predicted targets were included in pathway analysis if they were targeted by at least two microRNAs in the same regulatory direction (i.e. up or downregulated miRNAs in FMS).

The resulting target sets were subjected to pathway enrichment analysis using Reactome (Milacic et al., 2024), ShinyGo v.8 (Ge et al., 2020; Kanehisa et al., 2022; Luo and Brouwer, 2013), miRNA Tissue Atlas 2025 (Rishik et al., 2025) and EnrichR (Xie et al., 2021).

Statistical significance thresholds were defined as adjusted p-value of <0.05, after FDR correction, and only results that met this significance threshold were considered for further analysis.

## Results

### Literature search and identification of common dysregulated miRNAs in FMS

To identify miRNAs dysregulated in FMS across different cohorts, we searched published original research studies analyzing dysregulation of miRNAs using human clinical samples of patients with FMS (Figure 1). Fifteen primary research papers met our inclusion criteria (Table 1). These studies examined miRNA expression across different biofluids, such as blood, cerebrospinal fluid (CSF), and skin biopsies (Table 1). All but one study found statistically significant dysregulated microRNAs in FMS patients, either downregulated or upregulated, vs healthy controls. However, the overlap of differentially expressed miRNAs between studies was limited.

**Table 1:** List of primary research papers investigating dysregulation of microRNAs in fibromyalgia patients included in our study. MicroRNA levels were analyzed across different tissues in different cohorts. CSF: cerebrospinal fluid; PBMCs: peripheral mononuclear cells.

| **Tissues studied** | **Reference** | **DOI** |
| --- | --- | --- |
| Blood/Plasma/Serum, Skin | (Erbacher et al., 2025) | 10.1097/j.pain.0000000000003499 |
| Blood/Plasma/Serum | (Hernandez et al., 2024) | 10.1097/PR9.0000000000001199 |
| Blood/Plasma/Serum | (Erbacher et al., 2022) | 10.3390/cells11081276 |
| Blood/Plasma/Serum | (Akaslan et al., 2021) | 10.46497/ArchRheumatol.2022.8363 |
| Blood/Plasma/Serum | (Masotti et al., 2017) | 10.1007/s12035-016-0235-2 |
| Blood/Plasma/Serum | (Hussein et al., 2022) | 10.1093/pm/pnac076 |
| Blood/Plasma/Serum | (Nepotchatykh et al., 2023) | 10.1038/s41598-023-28955-9 |
| Blood/Plasma/Serum | (Bjersing et al., 2015) | 10.1007/s00296-014-3139-3 |
| Blood/Plasma/Serum | (Clos-Garcia et al., 2019) | 10.1016/j.ebiom.2019.07.031 |
| CSF | (Bjersing et al., 2013) | 10.1371/journal.pone.0078762 |
| Skin, PBMCs | (Leinders et al., 2016) | 10.1097/j.pain.0000000000000668 |
| PBMCs | (Braun et al., 2020) | 10.1371/journal.pone.0239286 |
| PBMCs | (Cerdá-Olmedo et al., 2015) | 10.1371/journal.pone.0121903 |
| PBMCs mitochondria | (Rasulova et al., 2025) | 10.1007/s11033-024-10110-w |
| Blood/Plasma/Serum | (Ayoub et al., 2025) | 10.1186/s40001-025-02330-y |

The majority of the studies aimed to find a distinct microRNA signature, and thus, for miRNAs to serve as clinical biomarkers of FMS disease. Since blood is easily collected, most studies used hematological (blood, serum or plasma) samples.

There was heterogeneity in the methodologies to extract RNA and analyze miRNAs, which can impact microRNA detection and quantification. Differences also included a wide range of patient samples sizes used per study. Reverse transcription coupled with quantitative polymerase chain reaction (RT-qPCR) was the most frequently used method for the analysis of miRNA levels (Table 2).

**Table 2:**
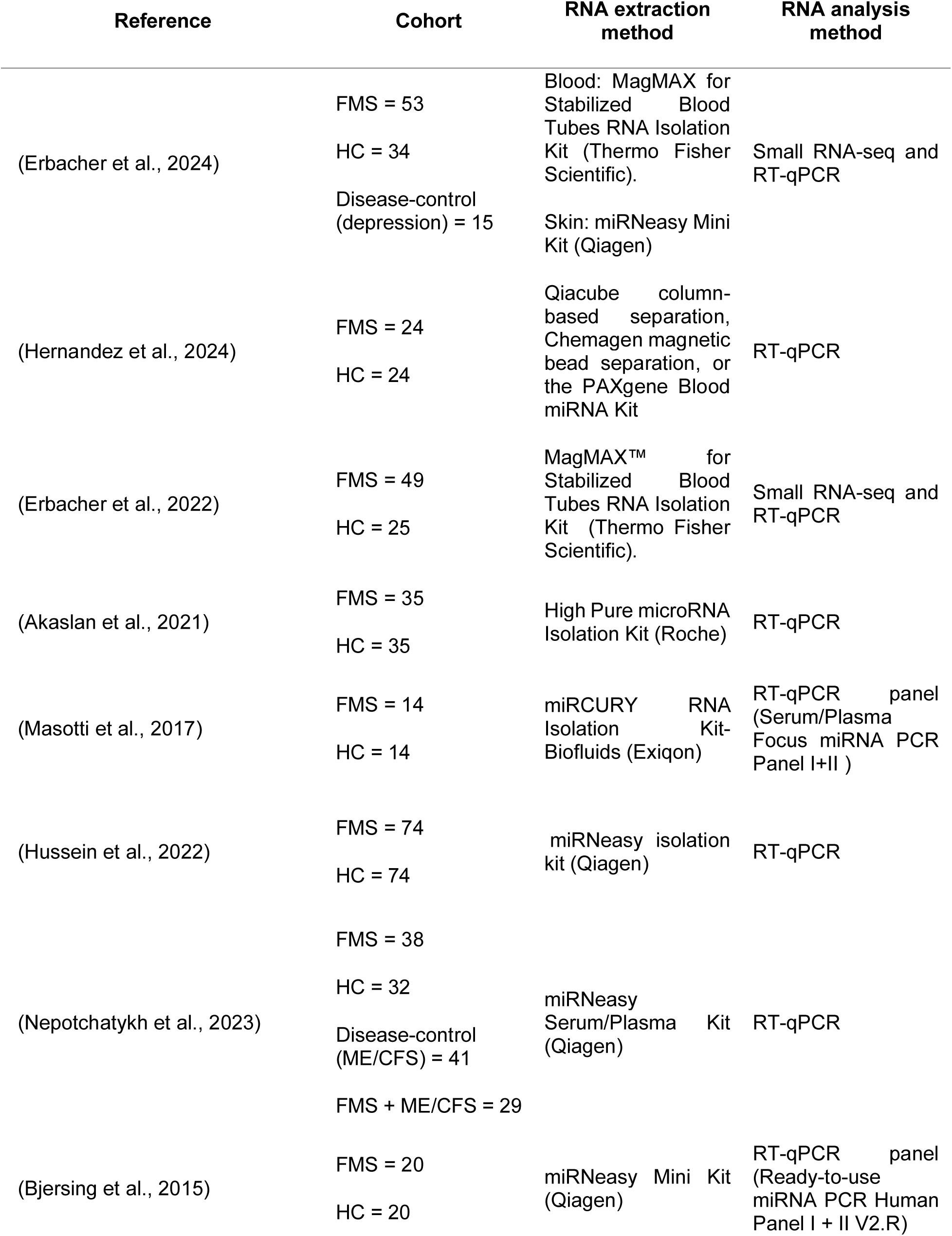
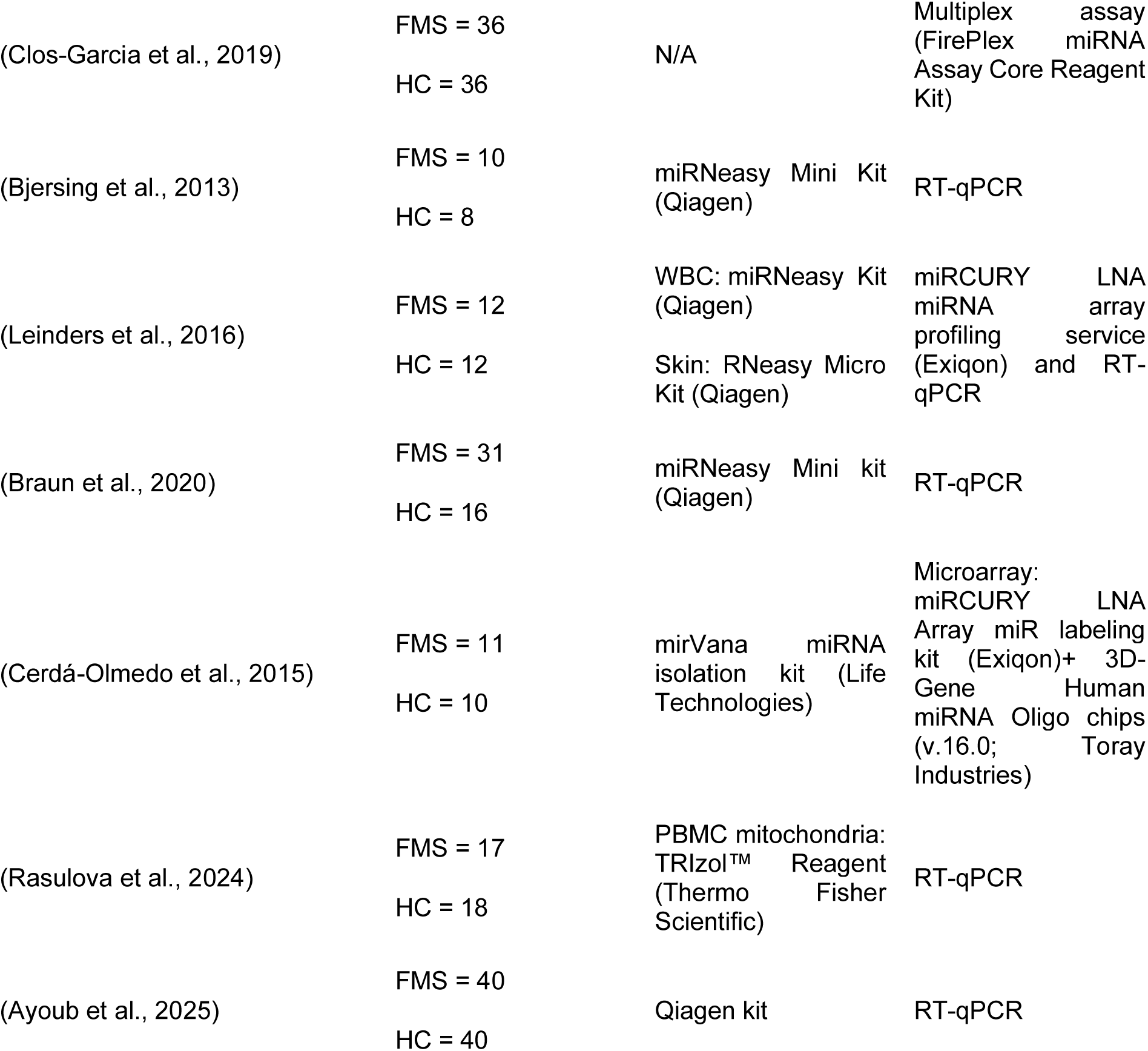
Cohort size and methodology for microRNA determination. Cohort size, RNA extraction method and miRNA levels determination method in our included studies.

Following our literature search, we extracted information regarding dysregulated microRNAs to assemble it into different working lists according to study, sample material, and direction of dysregulation. Different publications that used the same dataset were considered the same study in our analysis (Erbacher et al., 2024; Erbacher et al., 2022). Comparative analysis between the generated lists identified 10 miRNAs that were downregulated and 5 that were upregulated in at least two independent studies, irrespective of tissue (Table 3).

**Table 3:** Shared dysregulated microRNAs. List of shared microRNAs between at least two different studies.

| MicroRNA | Dysregulation direction | References |
| --- | --- | --- |
| hsa-miR-374b-5p | Downregulated | (Bjersing et al., 2015; Hernandez et al., 2024; Nepotchatykh et al., 2023) |
| hsa-miR-103a-3p | Downregulated | (Bjersing et al., 2015; Braun et al., 2020) |
| hsa-miR-107 | Downregulated | (Bjersing et al., 2015; Braun et al., 2020) |
| hsa-miR-127-3p | Downregulated | (Erbacher et al., 2024; Nepotchatykh et al., 2023) |
| hsa-miR-145-5p | Downregulated | (Bjersing et al., 2013; Cerdá-Olmedo et al., 2015) |
| hsa-miR-21-5p | Downregulated | (Bjersing et al., 2013; Cerdá-Olmedo et al., 2015) |
| hsa-miR-223-3p | Downregulated | (Bjersing et al., 2013; Cerdá-Olmedo et al., 2015) |
| hsa-miR-23a-3p | Downregulated | (Bjersing et al., 2013; Masotti et al., 2017) |
| hsa-miR-4433a-5p | Downregulated | (Erbacher et al., 2024; Nepotchatykh et al., 2023) |
| hsa-miR-532-3p | Downregulated | (Ayoub et al., 2025; Erbacher et al., 2024) |
| hsa-miR-320a | Upregulated | (Bjersing et al., 2015; Hussein et al., 2022) |
| hsa-miR-10a-5p | Upregulated | (Erbacher et al., 2024; Masotti et al., 2017) |
| hsa-miR-125a-5p | Upregulated | (Braun et al., 2020; Leinders et al., 2016) |
| hsa-miR-320b | Upregulated | (Hussein et al., 2022; Leinders et al., 2016) |
| hsa-miR-335-5p | Upregulated | (Clos-Garcia et al., 2019; Leinders et al., 2016) |

### Analysis of predicted biological effects of downregulated miRNAs

MiRNAs inhibit mRNA expression, and they can do so as networks where multiple microRNAs may target mRNAs that converge on specific biological functions (Hua et al., 2023). To infer possible biological pathways that the shared dysregulated miRNAs (Table 3) may influence, we predicted their mRNA targets using TargetScan v8 (McGeary et al., 2019), analysing up and downregulated miRNAs separately.

In total, 250 mRNA targets were predicted to be regulated by at least 2 downregulated miRNAs (Supplementary Table 1). and mapped onto biological pathways using Reactome, ShinyGO v0.80, and EnrichR(Ge et al., 2020; Milacic et al., 2024; Xie et al., 2021).

Enrichment analysis revealed that shared targets mapped to ‘Regulation of Phosphatase and TENsin homolog (PTEN) mRNA translation’ using Reactome (Figure 2A). KEGG database analysis identified the term ‘MicroRNAs in cancer’ as a result (not shown), while Gene Ontology (GO) biological process analysis using ShinyGO (Ge et al., 2020) indicated pathways related to RNA regulation, DNA transcription and development (Figure 2B).

**Figure 2.**
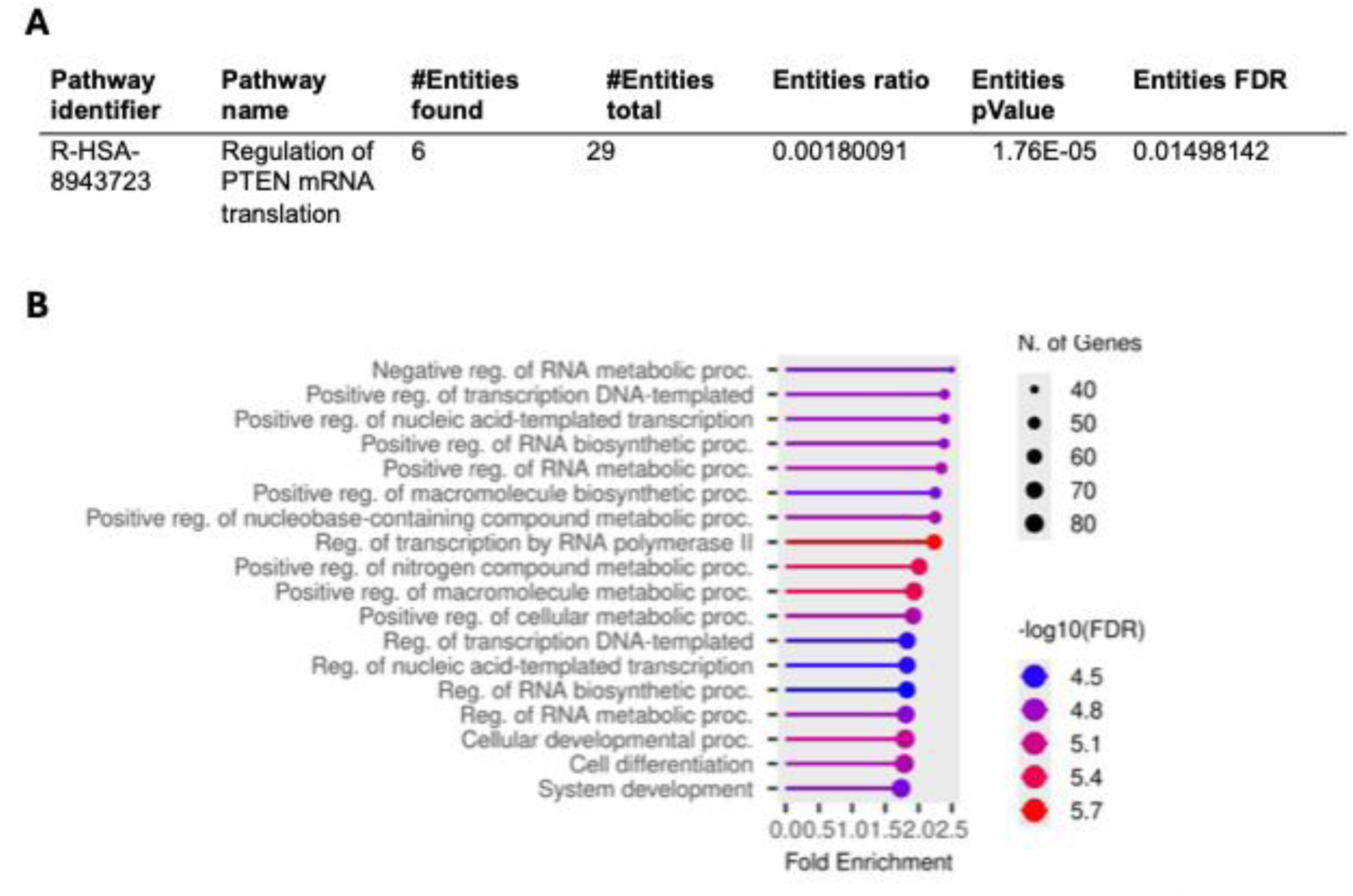
Prediction of Biological Effects of Downregulated microRNAs. A. Reactome results. ‘PTEN mRNA translation’ was identified as biological pathway to which the common targets of downregulated microRNAs in FMS mapped. B. Fold enrichment plot of GO Biological Processes identified by ShinyGo V8. Multiple pathways related to RNA metabolism were predicted to contain targets of commonly downregulated microRNAs in FMS.

Furthermore, four mRNA targets were predicted to be regulated by 4 out of 10 of the downregulated miRNAs (Table 4), suggesting potential convergence in the regulation of expression of specific genes. These mRNA targets encode proteins that relate to multiple cellular processes, including an acetyltransferase (*NAA50*), protein transport (*SNX24*), iron uptake (*TFRC*) or TGF-β binding (*TGFBR2*) (https://www.genecards.org).

**Table 4:**
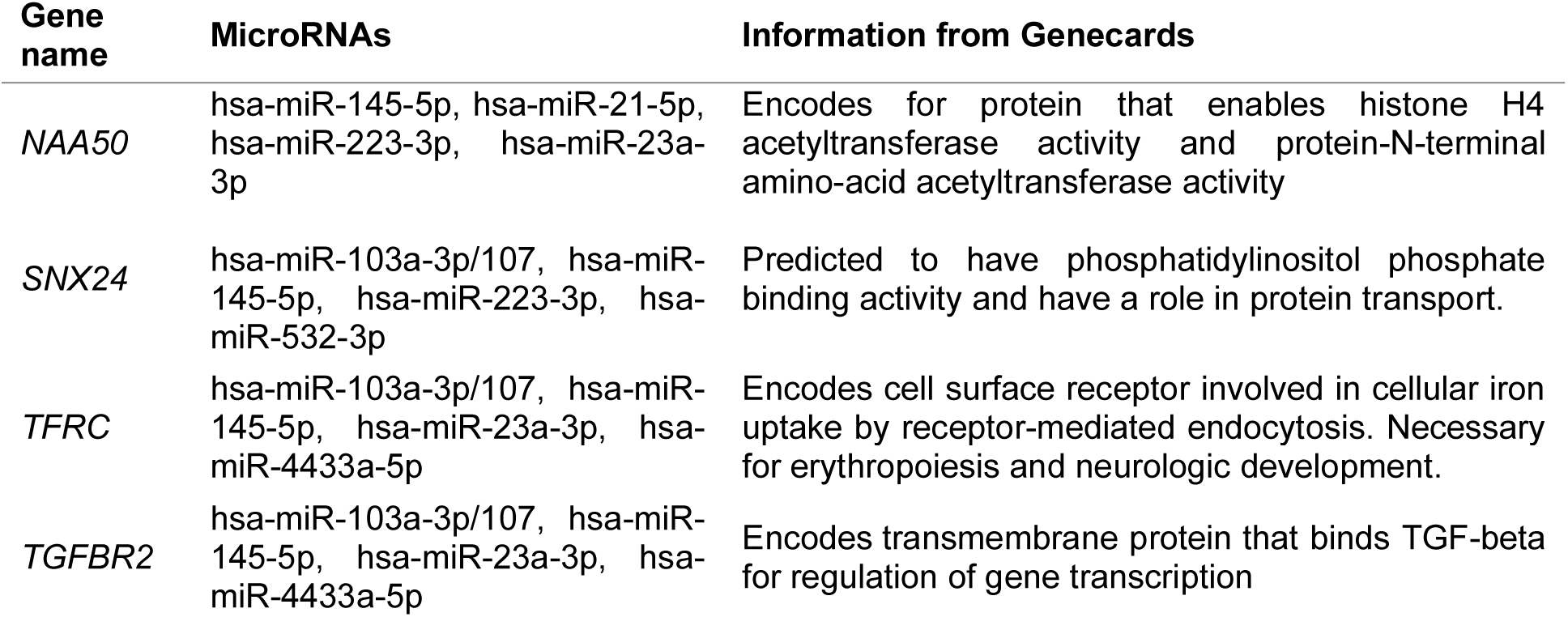
mRNAs predicted to be targeted by at least 4 microRNAs downregulated in FMS. Information of the target genes was obtained from GeneCards Human Gene Database (https://www.genecards.org).

### Analysis of predicted biological effects of upregulated microRNAs

The five commonly upregulated miRNAs were analyzed using the same method. Their predicted targets were 46 mRNAs (Supplementary Table 2).

Reactome pathway enrichment mapped to ‘Nuclear Receptor Transcription’ (Figure 3A), while KEGG analysis (Ge et al., 2020; Kanehisa et al., 2022) predicted an altered circadian rhythm (Supplementary Figure 1). GO Biological processes related to metabolic processes, and nucleic acid-related pathways were also highlighted (Figure 3B).

**Figure 3.**
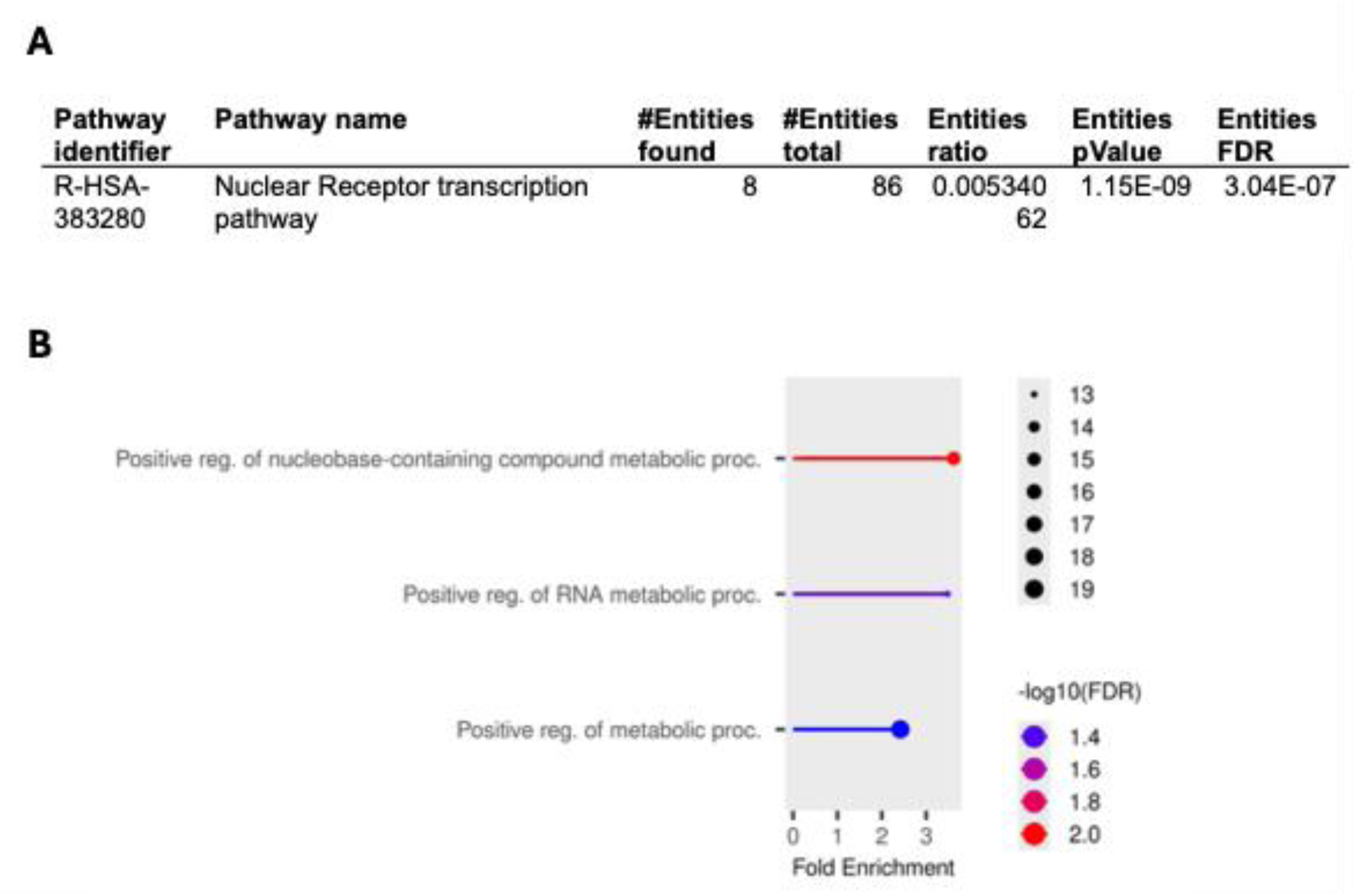
Prediction of Biological Effects of Upregulated microRNAs. A. Reactome results. ‘Nuclear receptor transcription pathway’ was identified as biological pathway to which the common targets of upregulated microRNAs in FMS mapped. B. Fold enrichment plot of GO Biological Processes identified by ShinyGo V8. Multiple pathways related to RNA metabolism were predicted to contain targets of commonly upregulated microRNAs in FMS.

### Integrated Analysis of Upregulated and Downregulated microRNA Targets

As miRNAs can act within complex regulatory networks, mRNAs targeted by both upregulated and downregulated miRNAs can converge into similar biological pathways. Thus, we additionally performed an integrated analysis combining the targets of both sets of dysregulated microRNAs to identify if there were pathways implicated in this manner.

Reactome and ShinyGO BP pathways (Figures 4A and 4B) were similar to results from the predicted targets by downregulated miRNAs (Figure 2). EnrichR identified targets significantly different in Type I Spiral Ganglion Neuron Brain Mouse (Figure 4C).

**Figure 4.**
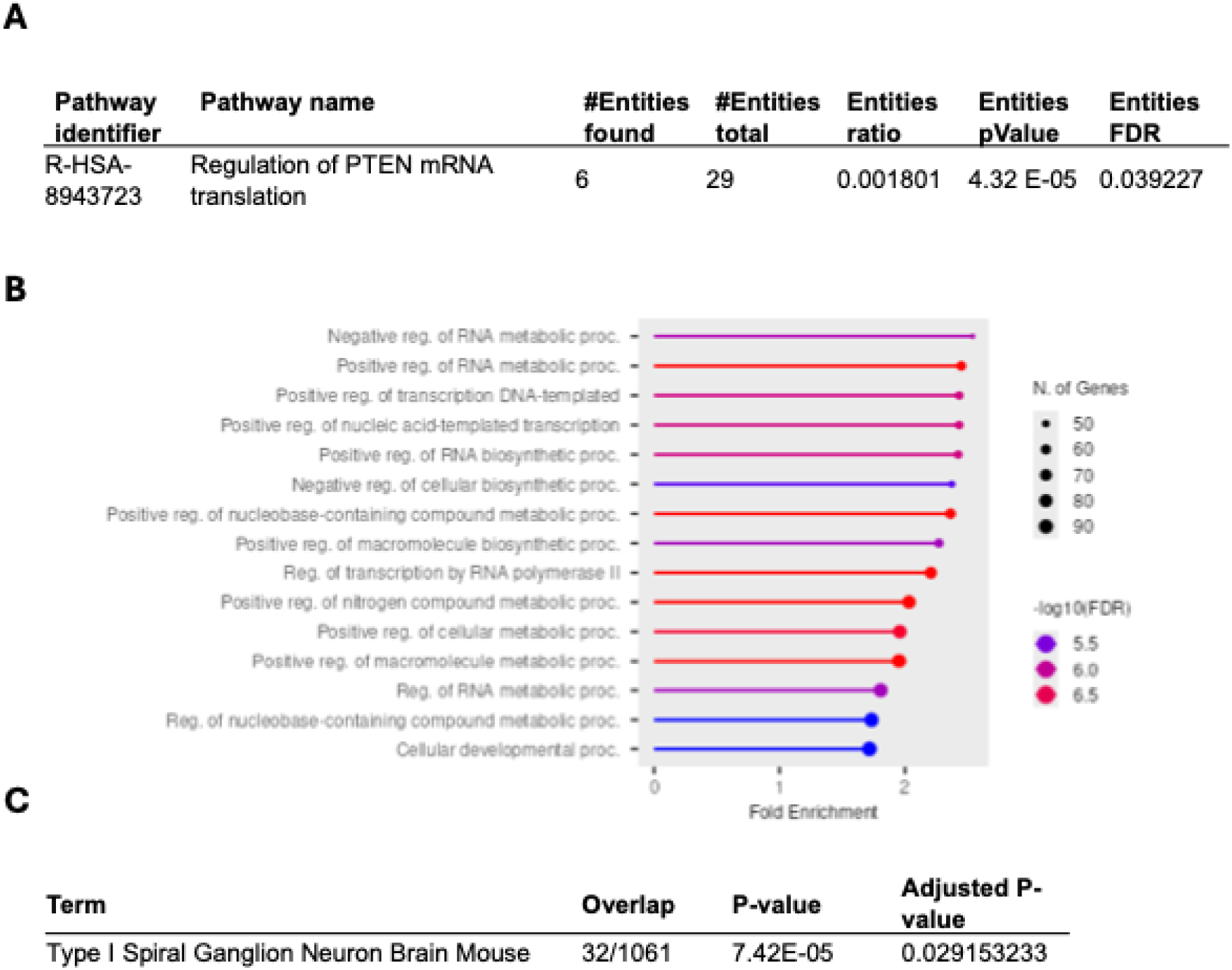
Prediction of Biological Effects of Up and Downregulated microRNAs. A. Reactome results. ‘PTEN mRNA translation’ was identified as biological pathway to which the targets of upregulated and downregulated microRNAs in FMS mapped. B. Fold enrichment plot of GO Biological Processes identified by ShinyGo V8. C. Type I Spiral Ganglion Neuron Brain Mouse identified by EnrichR.

### Tissue Distribution of Dysregulated miRNAs

So far, our analysis suggested that dysregulated microRNAs in FMS may alter fundamental cellular processes including nucleic acid metabolism and neuronal pathways. Since the majority of analyzed studies investigated circulating miRNAs, their origin (or their end-point targeting) and thus their tissue distribution remained unclear. To infer potential tissue associations, we utilized microRNA Tissue Atlas 2025 (Rishik et al., 2025). We were able to locate the top 15 tissues of the detected as commonly downregulated and upregulated miRNAs in FMS (Table 3). We then annotated the top tissues that were shared by at least half of the miRNAs in each group (up or downregulated) (Figure 5).

**Figure 5:**
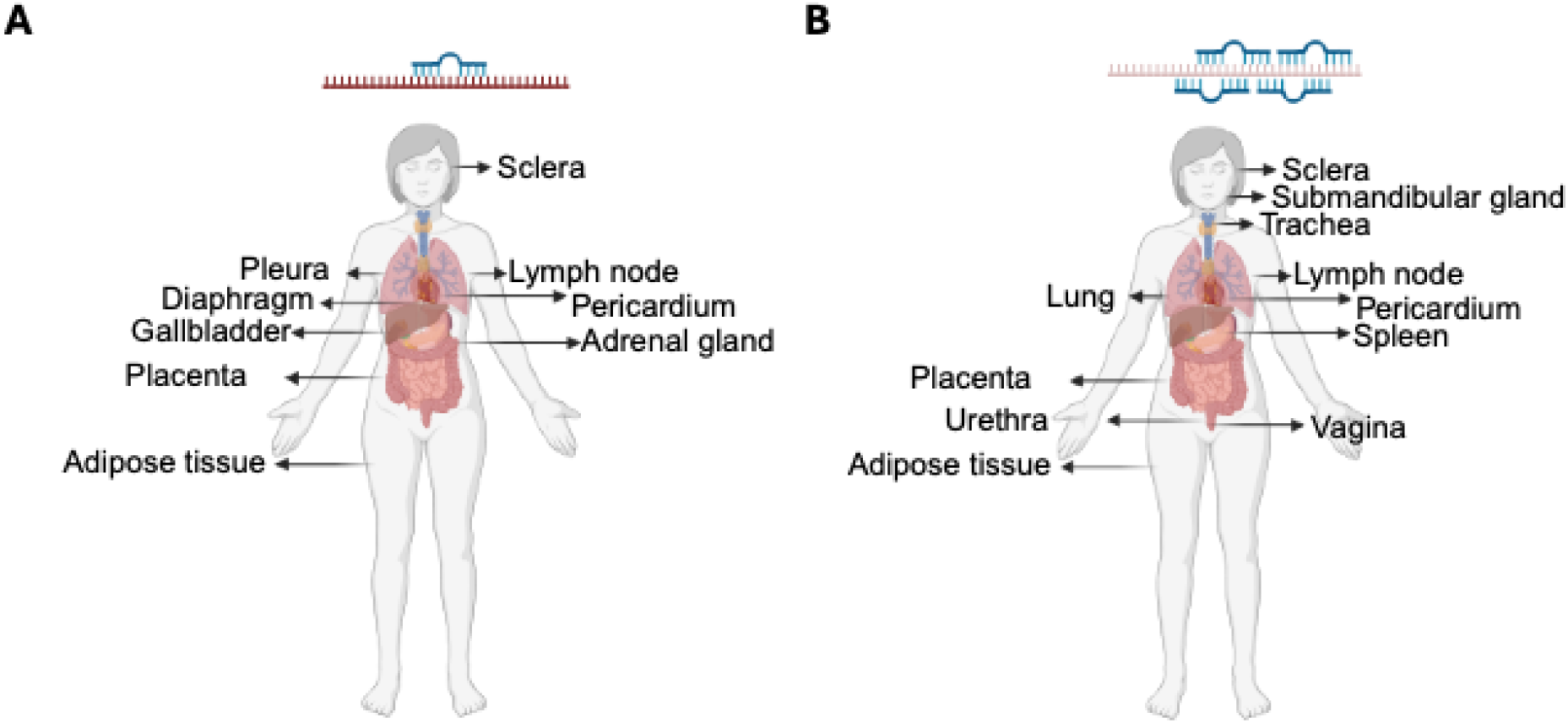
Tissue localization of microRNAs dysregulated in FMS. Top 10 tissues where dysregulated microRNAs in FMS were found. A. Localization of downregulated microRNAs in FMS vs healthy. B. Localization of upregulated miRNAs in FMS vs healthy. Created in BioRender. Martinez-nunez, R. T. (2026) https://BioRender.com/cwk6ww0.

We found 11 top tissue locations shared by at least five downregulated miRNAs (Figure 5A) and 9 top tissue locations shared by at least three upregulated miRNAs (Figure 5B). There was a wide variety of tissues including adrenal gland, diaphragm, trachea or sclera. Both upregulated and downregulated miRNAs mapped to adipose, lymph node, pericardium, placenta and sclera, suggesting an important role for these tissues, as origin or target, of the detected dysregulated microRNA in FMS. These observations are consistent with the notion that FMS affects multiple organ systems, and that it is not limited to the nervous system.

## Discussion

FMS is a complex disorder with diverse clinical manifestations and symptoms that affect multiple organs, presenting as widespread pain, fatigue and cognitive problems. Circulating microRNAs are a potential vehicle to (dys)regulate processes across tissues and have been evaluated in multiple FMS cohorts. To determine a common microRNA signature in FMS and infer its possible pathophysiological role, we analyzed previous literature that assessed dysregulated microRNA levels in FMS. Across 14 original manuscripts, we analyzed the potential role of microRNAs that were dysregulated in the same direction (up or down) in FMS patients, in at least two independent cohorts. We found hsa-miR-320a, hsa-miR-10a-5p, hsa-miR-125a-5p, hsa-miR-320b and hsa-miR-335-5p upregulated and hsa-miR-374b-5p, hsa-miR-103a-3p, hsa-miR-107, hsa-mir-127-3p, hsa-miR-145-5p, hsa-miR-21-5p, hsa-miR-223-3p, hsa-miR-23a-3p, hsa-miR-4433a-5p and hsa-miR-532-3p downregulated. The predicted targets of these miRNAs mapped to immunological pathways, such as regulation of PTEN (Tsai et al., 2024), or neurological pathways such as type I spiral ganglion neurons (Figure 4). Additionally, we found that dysregulated microRNAs were over-represented in a diverse range of tissues (Rishik et al., 2025), including lymph node, adipose tissue and spleen. Overall, our analysis supports dysregulation of microRNAs as a mechanism that may affect multiple tissues by modulation of neuroimmune and ubiquitous pathways in FMS patients.

Downregulated microRNAs were predicted to target genes (*AGO1*, *PTEN* and *CNOT6L*) that mapped to ‘regulation of PTEN mRNA translation’ (Figure 2). *PTEN* is a known tumour suppressor gene that regulates cell growth and proliferation (Taylor et al., 2019) through the phosphatidylinositol 3-kinase (PI3K)/protein kinase B (PI3K/AKT) and mTOR signaling pathways (Zhou et al., 2024). Patients with mutations that disrupt PTEN activity can develop autoimmunity, lymphoid hyperplasia (Chen et al., 2017), and inflammation due to activated B cells (Tsai et al., 2024), possibly affecting peripheral immune tolerance (Jaini et al., 2020). Furthermore, Gene Ontology (GO) biological process analysis also supported the possibility of development and differentiation of cells being affected (Figure 2B), concomitant with PTEN pathways being dysregulated (Taylor et al., 2019).

PTEN has previously been linked to pain modulation, although its precise role remains to be explored in detail. On the one hand, upregulation of PTEN ameliorated hypersensitivities in the sciatic nerve chronic constriction injury model of neuropathic pain (Fang et al., 2022; Huang et al., 2015), but on the other, shRNA-mediated knockdown of PTEN reduced allodynia in a rat model of trigeminal neuropathy (Li et al., 2019). Although there is no direct evidence available for a role in FMS, the possibility that dysregulation of PTEN is present in patients deserves to be explored.

Our analysis also revealed that four transcripts (*NAA50*, *SNX24*, *TFRC*, *TGFBR2*) were targeted by more than 4 downregulated microRNAs (Table 4). Because miRNAs can act as networks, multiple microRNAs converging on specific mRNA species strongly indicates a physiologically relevant mRNA regulation (An et al., 2010). The number of targets identified here was too small to perform a meaningful enrichment analysis, but individually they could suggest immune dysregulation via dysregulation of TGF-β action (*TGFBR2* encodes for a TGF-β receptor subtype), a main mediator of wound healing and immune activation (Gilbert et al., 2016). *TFRC*, which encodes for transferrin receptor 1 (TfR1), is involved in iron uptake and development of neuronal cells (Stelzer et al., 2016), with particular importance for the nervous and haematopoietic systems (Levy et al., 1999). Reduced iron availability in lymphocytes leads to impaired function, and patients with mutations in the *TFRC* gene have been linked to immunodeficiency and mitochondrial impairment (Aljohani, 2026; Muñoz-Pujol G, 2026).

Many FMS patients report sleep disruption and an altered circadian rhythm (Mahdi et al., 2011). Sleep governs multiple immune functions and sleep disruption can compromise the immune system (Szredzka et al., 2025). Circadian rhythm was predicted to be altered by upregulated microRNAs (Fig 2). Dysregulation of circadian rhythm can also lead to metabolic changes. Indeed, GO analysis indicate changes in metabolic processes, where some of the mapped targets (such as *PRKAA2*, *MTF1*, *TRIAP1,* Figure 2) are related to metabolic, cellular, and mitochondrial stress (Stelzer et al., 2016). Moreover, mitochondrial dysfunction in PBMCs can be present in FMS patients (Israel et al., 2024), which has been suggested to contribute to the fatigue that characterizes the condition (Macchi et al., 2024).

In addition to associations with diagnosis, some investigations also indicate a relationship between miRNA dysregulation and symptom severity, and others suggest potential links to processes such as glutamate metabolism, mitochondrial dysfunction, nociceptive fiber alterations, and immune-related pathways (Bjersing et al., 2015; Braun et al., 2020; Cerdá-Olmedo et al., 2015; Clos-Garcia et al., 2019; Erbacher et al., 2022; Leinders et al., 2016; Rasulova et al., 2024).

Our analysis identified multiple core cellular processes as potentially targeted by miRNA dysregulation. These included nuclear receptor transcription pathway (Figure 3), vital for immune-inflammatory responses to signals from hormone and lipid mediators (Jin et al., 2025). We also found core RNA processing and nucleic acid processing pathways (Figure 3), which may indicate widespread dysregulation of basic cellular functions that relate to cell proliferation or differentiation, important for homeostasis.

In keeping with the larger number of downregulated miRNAs (10) and corresponding targets (250), compared to upregulated miRNAs (5) and their targets (46), the pathways potentially altered by both up- or downregulated miRNAs in FMS were largely driven by the downregulated miRNAs (Figure 2). Consequently, analyzing all targets together identified similar pathways to those that downregulated microRNAs mapped to (Figure 2 and 4). Cell Type I spiral ganglion neuron (Brain from mouse) was identified as a new pathway in our combined analysis (Figure 4). These cells are the main type of primary sensory neurons in the cochlea, part of the auditory system, transmitting auditory information to the brain (Grandi et al., 2020). Potentially, this observation may be relevant to the hearing problems regularly reported by FMS patients (Tuncer et al., 2021). Our combined analysis also showed an over-representation of targets mapping to RNA and nucleic acid metabolic processes (Figure 4) that overshadowed other pathways from enrichment analysis when investigating only upregulated microRNAs. These data highlight the value of breaking down the analysis to further understand the possible role of microRNAs, and the potential mechanisms behind. Our analysis also highlights that there appears to be a widespread downregulation of microRNAs in FMS that renders wider effects (250 targets/10 microRNAs) than those upregulated (46/5 microRNAs). This difference may indicate a generalized deficient mechanism of microRNA cargo in FMS.

FMS is a complex disorder with multisystemic manifestations. Accordingly, dysregulated miRNAs appeared to be over-represented in tissues from different parts of the body (Figure 5). Our analysis cannot discern if those tissues may be affected by the miRNA dysregulation or if they may be the source of dysregulated microRNAs. Several of the implicated tissues are related to immune function, such as lymph node, spleen and adrenal gland (Figure 5), which are in accordance with the immune dysfunction observations found in patients with FMS. The identification of adipose tissue may be related to metabolic function (An et al., 2023); but is also involved in secretion of inflammatory and immune factors in FMS (Gerdle et al., 2024). Furthermore, FMS patients commonly have a higher BMI (Atzeni et al., 2021).

Here, we have performed a comprehensive characterization of common microRNA signatures in FMS. However, we acknowledge that our study comes with limitations. We can confidently say that the microRNAs we investigated were consistently found dysregulated across different cohorts and are likely to be a consequence, and possibly contribute to severity of FMS. Our findings are based on *in silico* predictions rather than molecular validations, but on the other hand, employ multiple independent datasets. To minimize false miRNA:target predictions, we used the gold-standard prediction tool TargetScan (McGeary et al., 2019) for microRNA-target prediction. TargetScan relies on multiple parameters, including conservation amongst species, that makes it more restrictive than other tools (Quillet et al., 2021). Furthermore, while several microRNAs were found to be significantly dysregulated in individual studies of FMS patients, only a small number of them were replicated in independent studies. This may be due to different processing methods and analyses, but also due to heterogeneity of the disease and clinical characteristics of the different patient cohorts. By restricting our analyses to microRNAs that were found dysregulated in 2 or more studies, we believe that the set of microRNAs presented here may be more robust candidates in FMS.

Future investigations may shed light on the consistency of the dysregulated microRNA signature in larger cohorts. The field would benefit from protocol harmonization for sample processing and an increased emphasis on RNA sequencing rather than qPCR/microarrays, as the former is an unbiased approach (Benesova et al., 2021). Investigations of miRNAs in extracellular vesicles is an attractive future analytical route that should be explored. Since extracellular vesicles provide a more protective environment compared to freely circulating miRNAs, they may contain miRNAs that are more likely to have functional consequences (He et al., 2021) and that may be more reproducibly found in different cohorts or potential diagnostic use.

Molecular studies would shed light on the role of the targets inferred. Investigations of PTEN related pathways, nuclear receptors and cellular metabolic stress in the context of FMS could reveal mechanistic insights and advance our understanding of the condition. For example, PTEN modulation is currently being investigated in oncology and neurology (McLoughlin et al., 2018). Elucidating the cellular origins of the dysregulated miRNAs and their targets is very likely to advance our understanding of FMS pathophysiology. Importantly, the pattern of miRNA dysregulation identified by our analysis strongly indicates that FMS is a systemic disorder rather than a condition defined by aberrant cortical processing.

In summary, we present an analysis of dysregulated miRNAs in FMS. We found 10 miRNAs commonly downregulated and 5 miRNAs commonly upregulated in FMS vs healthy, pain-free subjects. These microRNAs map to PTEN regulation pathways, nuclear receptor transcription pathways and metabolic processes as possibly dysregulated and correlated to FMS pathogenesis. Our data warrants further research to both validate predicted data and investigate the biological significance of miRNA dysregulation in FMS.

## Data Availability

All data produced in the present work are contained in the manuscript

## Acknowledgements

ATO was funded by the Wellcome Trust Neuro-Immune Interactions in Health and Disease PhD Program (218452/Z/19/Z).

## Author contributions

ATO: Data curation, formal analysis, funding acquisition, investigation, methodology, writing – original draft, writing – review and editing. DA and RTMN: Conceptualization, funding acquisition, investigation, methodology, supervision, writing – original draft, writing – review and editing.

## Disclosures

RTMN declares speaker honoraria from AstraZeneca and GSK not related with the subject in this manuscript.

## Notes

### Author Declarations

DOIs: 10.1097/j.pain.0000000000003499 10.1097/PR9.0000000000001199 10.3390/cells11081276 10.46497/ArchRheumatol.2022.8363 10.1007/s12035-016-0235-2 10.1093/pm/pnac076 10.1038/s41598-023-28955-9 10.1007/s00296-014-3139-3 10.1016/j.ebiom.2019.07.031 10.1371/journal.pone.0078762 10.1097/j.pain.0000000000000668 10.1371/journal.pone.0239286 10.1371/journal.pone.0121903 10.1007/s11033-024-10110-w 10.1186/s40001-025-02330-y

