## Supplementary figures and images for "Dysregulation of microRNAs in fibromyalgia: potential as biomarkers and disorder modulators"

### Supplementary Figure 1

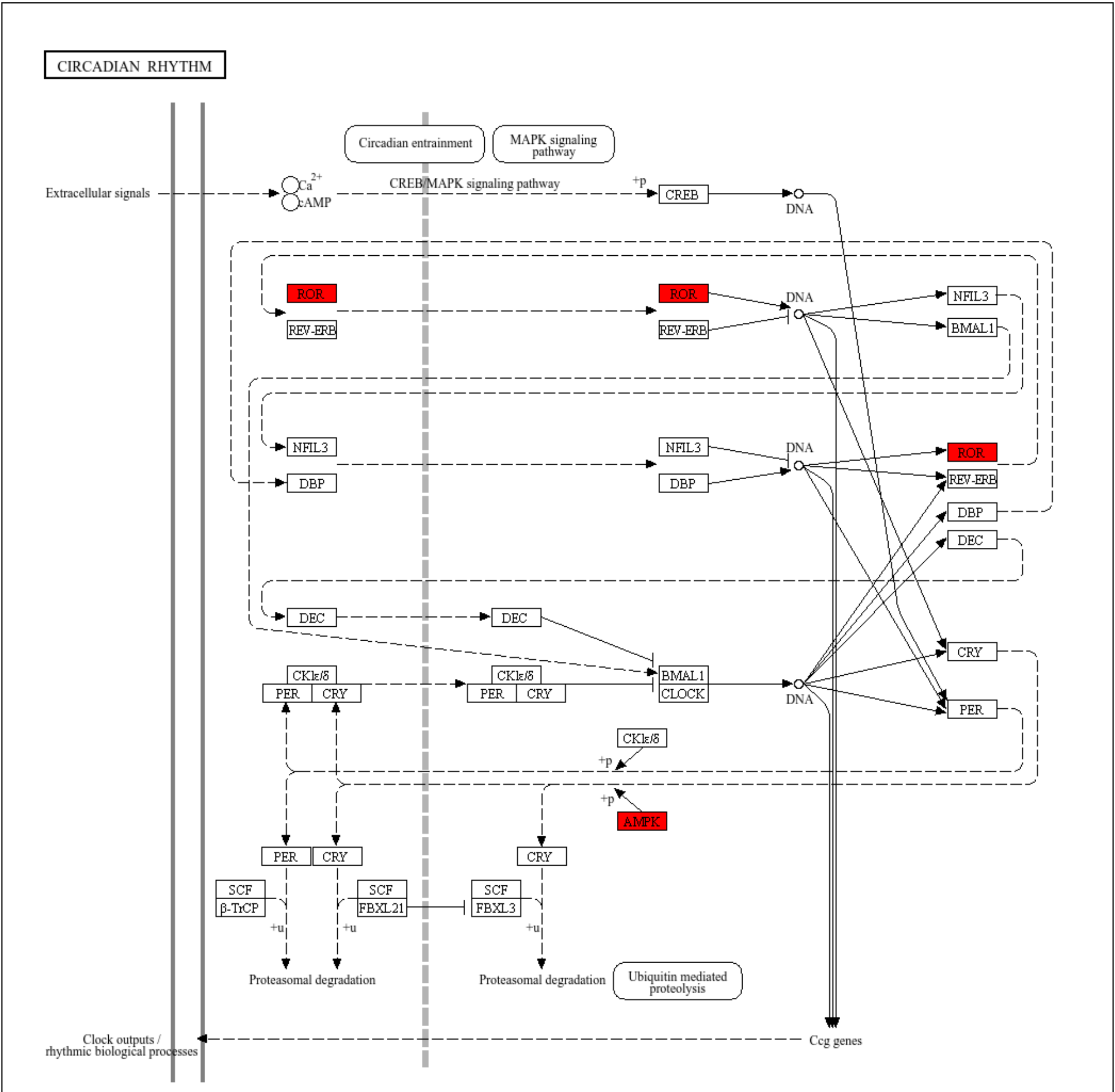


Data on KEGG graph
Rendered by Pathview
